# Obesity paradox in patients with cancer: A systematic review and meta-analysis of 6,320,365 patients

**DOI:** 10.1101/2020.04.28.20082800

**Authors:** Fausto Petrelli, Alessio Cortellini, Alice Indini, Gianluca Tomasello, Michele Ghidini, Olga Nigro, Massimiliano Salati, Lorenzo Dottorini, Alessandro Iaculli, Antonio Varricchio, Valentina Rampulla, Sandro Barni, Mary Cabiddu, Antonio Bossi, Antonio Ghidini, Alberto Zaniboni

## Abstract

**BACKGROUND:** Obesity, defined as a body mass index (BMI) > 30 kg/m2, is associated with a significant increase in risk of many cancers. In last years, various studies suggested that obese cancer patients have better outcomes than non-obese patients. This phenomenon, also known as “the obesity paradox”, is not well understood and presents controversial explanations. We performed a systematic review and meta-analysis to assess the association between obesity and outcome after a diagnosis of cancer.

**PATIENTS AND METHODS:** PubMed, the Cochrane Library, and EMBASE were searched from inception to January 2020, for studies reporting prognosis of patients with obesity and cancer. Risk of death, cancer-specific survival (CSS) and progression were pooled to provide an adjusted hazard ratio with a 95% confidence interval (HR 95%CI). The primary outcome of the study refers to overall survival (OS) in obese vs non-obese patients with malignancies. Secondary endpoints were CSS and progression- or disease-free survival (PFS or RFS).

**RESULTS:** Mortality and relapse associated with obesity in patients with cancer were evaluated among n=6,320,365 participants (n=203 studies). Overall, association of obesity and cancer was associated with a reduced OS (HR =1.14, 95% CI: 1.09-1.19; P<.01) and CSS (HR=1.17, 95%CI 1.12-1.23; P<.01). Patients were also at increased risk for relapse (HR=1.13, 95%CI 1.07-1.19; P<.01). Patients with breast, colorectal and uterine tumors were at increased risk of death. Conversely, obese with lung cancer, renal cell carcinoma and melanoma survived longer that non-obese.

**CONCLUSIONS:** In many cancer patients, obesity reduces survival and increases the risk of relapse. In lung cancer, renal cell carcinoma and melanoma obesity was protective in terms of outcome. More intensive follow up, adequate dosing of oncological treatments, calories intake restrictions, physical activity and monitoring of obesity-related complications are effective measures for reducing mortality in these subjects.

## Introduction

Obesity, defined as a body mass index (BMI) greater than 30 kg/m2, is a chronic disease with an increasing prevalence around the world, largely contributing to important health issues in most countries. Despite body fat is a general risk factor for serious illness (e.g. metabolic syndrome), greater cardiometabolic risk has also been associated with the localization of excess fat in the visceral adipose tissue and ectopic deposits^1^. Several large epidemiologic studies have evaluated the link between obesity and mortality. In particular, a meta-analysis of 230 cohort studies including over 30 million individuals, found that both obesity and overweight were associated with an increased risk of all-cause mortality^2^. Despite the evidence that excess mortality increases with increasing BMI, some studies have reached the conclusion that elevated BMI may improve survival in subjects with cardiovascular disease, a phenomenon called the “obesity paradox”^3^.

Increased BMI is also associated with an increased risk of multiple cancer types^4^. In addition, obesity and overweight may increase cancer mortality^5^. During last decades we also observed a more rapid increase in obesity among adult cancer survivors compared with the general population^6^. The mechanisms contributing to higher cancer incidence and mortality may include alterations in sex hormone metabolism, insulin and insulin-like growth factor levels, and adipokine pathways^7 8^.

In last years, various studies suggested that, in cancers, patients with a normal BMI have worse outcomes than obese patients. This phenomenon (the obesity paradox) in cancer, is not well understood and presents controversial explanations^9 10 11^. Three different metaanalyses led to different results in particular in lung and renal cell carcinomas^12 13 14^. In lung cancer, obesity has favourable effects on long-term survival of surgical patients. Moreover, in renal cell carcinoma an inconsistent effect of BMI on cancer-specific survival was found. Conversely, breast, ovarian and colorectal cancer are invariably associated with increased mortality in obese patients^15 16 17^. The main explanations proposed to answer these observations include the general poor health status of patients with very low BMI. Additionally, weight loss may usually be associated with frailty and other risk factors (e.g. smoke)^10^. In cancer, obesity is also associated with increased efficacy of PD-1/PD-L1 blockade in both tumor-bearing mice and patients^11^.

This updated systematic review and meta-analysis was conducted to evaluate the prognosis of obese vs non-obese cancer patients.

## Material and methods

We followed the Preferred Reporting Items for Systematic Reviews and Meta-analyses (PRISMA) and Meta-analysis of Observational Studies in Epidemiology (MOOSE) reporting guidelines^18 19^.

### Search strategy and inclusion criteria

A systematic search was conducted of the EMBASE, Pubmed and The Cochrane Library published from inception until January 2020. The following search terms were used: *((carcinoma or cancer or sarcoma or melanoma or (“Neoplasms”[Mesh])) AND (obese OR obesity OR 30 kg/m*^2^ *OR “body mass index”) AND (hazard ratio) AND survival AND (multivariate OR cox)*. The reference lists of identified articles were then manually searched to identify potentially relevant omitted citations Articles that were not published in the English language were not included in this study.

Retrospective and observational studies (cohort, case control) or prospective trials were selected when they reported the association of obesity, defined as a BMI >30 kg/m2, with the risk of death (OS), cancer-specific death (CSS) or disease-progression (PFS or DFS) in patients with cancer. We placed no restrictions on study setting, size, race or country. The inclusion was limited to studies reporting hazard ratios (HRs) and their corresponding 95% CIs. Studies were restricted to adult patients with solid tumors. Hematologic malignancies were excluded.

The most up-to-date versions of full-text publications were included. Study selection was performed in 2 stages, first, titles and abstracts were screened; then, selected full-text articles were included according to the eligibility criteria. Screening was performed independently by 10 authors (MG, GT, AG, AI, AC, ON, VR, AI, LD, MS) and conflicts were handled by consensus with a senior author (FP).

### Data Collection and quality assessment

Data were collected independently by using a predesigned spreadsheet (Excel [Microsoft]. Collected data items included authors, year of publication, study setting and design, median follow up, treatments received, outcomes, and type of analysis. The primary outcome was OS, secondary endpoints were CSS and PFS/DFS. Along with data extraction, 1 author (FP) assessed study quality according to a modified Newcastle Ottawa Scale^20^.

### Statistical analysis

First, pooled HRs with 95% CIs were estimated using random effects meta-analysis with the generic inverse-variance method for studies that provided fully adjusted HRs. Inconsistency across studies was measured with the I2 method. Cutoff values of 25%, 50%, and 75% indicated low, moderate, and high heterogeneity, respectively. When I2 was larger than 50%, a random effects model was used; otherwise, the fixed effects model was used. Second, to examine heterogeneity, we performed analyses of predefined subgroups based on type of disease. Additionally, to address potential bias and verify our results, we performed sensitivity analysis using a leave-one-out method and the Trim and Fill method. Finally, to investigate the risk of publication bias, we applied the Egger test and visually inspected the funnel plots (begg’s test). All analyses were carried out using Comprehensive Meta-Analysis software,

## Results

Our literature search yielded 1,892 articles, of which 203 met the inclusion criteria for our overall systematic review of the effect of obesity on cancer outcome^21,22,31,121–130,32,131–140,33,141–150,34,151–160,35,161–170,36,171–180,37,181–190,38,191–200,39,201–210,40,211–220,23,41,221–223,42–50,24,51–60,25,61–70,26,71–80,27,81–90,28,91–100,29,101–110,30,111–120^ (Fig. 1). Most excluded studies did not use the cut-off value for obesity (BMI values different from 30 kg/m2) or used different cut-off for risk of death (e.g. 1 kg/m2 of increase in BMI). Of the 203 articles, 170 were eligible for inclusion in the systematic review of the risk of obesity with OS, 109 for correlation with CSS, and 79 for correlation with DFS/PFS. Descriptive data for studies included in our meta-analysis are listed in Table 1. Overall, the included studies encompassed a total of 6,320,365 patients. Sample sizes ranged from 41 to 1,096,492 with a median of 1,543. Most studies were retrospective in nature (n=132), the minority were prospective cohort/observational studies (n=63) or pooled analysis/randomized trials (n=8).

**Fig. 1.**
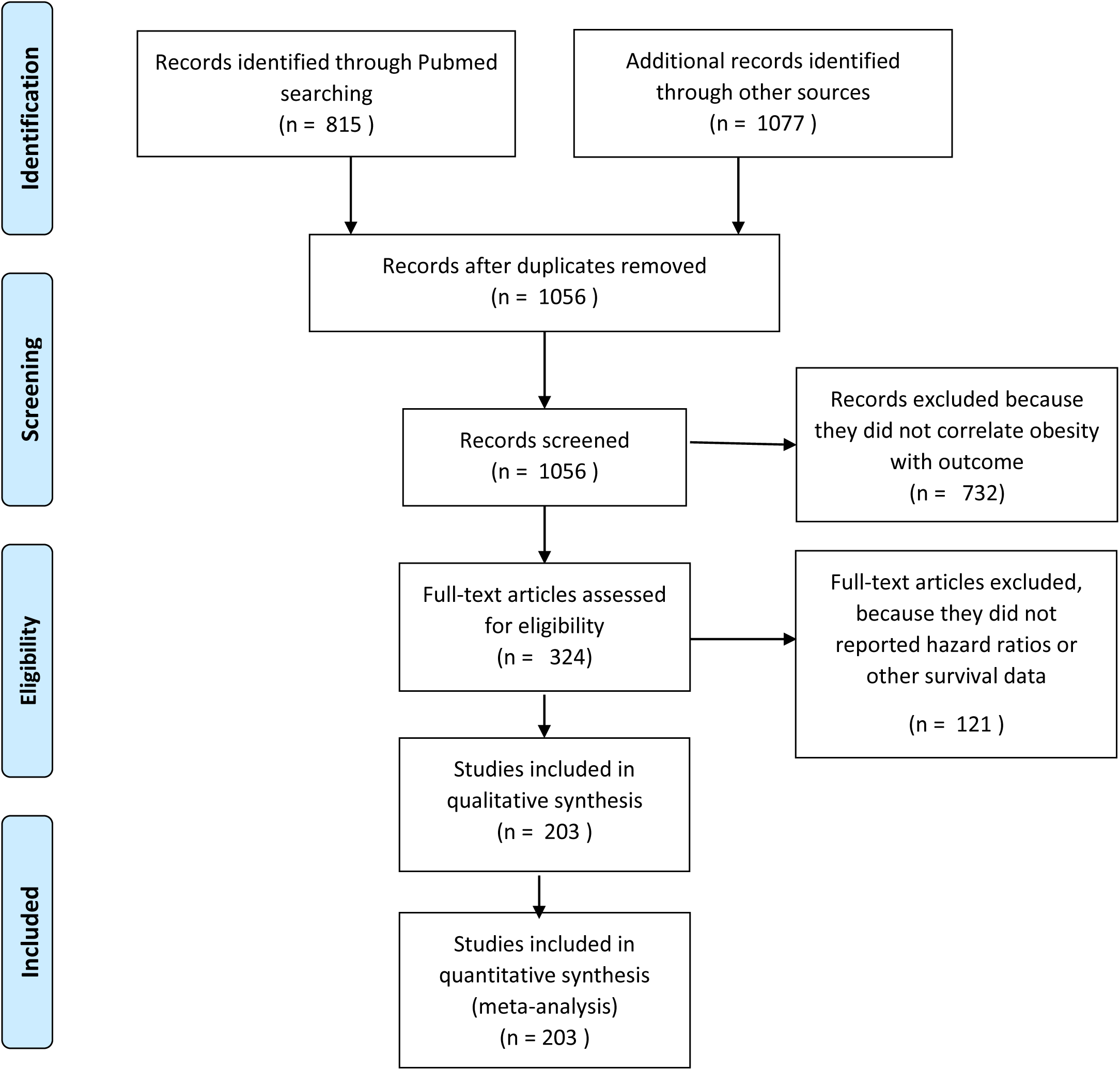
flow diagram of included studies.

**Tab. 1:** characteristics of included studies.

| Author, Year | N° pts | N° of obese (%) | Study type | Country | Disease | Median Follow Up (months) | Disease stage | Treatment | Type of analysis | OS | CSS | DFS/PFS | Quality (NOS-METRICS) |
| --- | --- | --- | --- | --- | --- | --- | --- | --- | --- | --- | --- | --- | --- |
| Abdel-Rahman/2019<sup>21</sup> | 145544 | 18131 (23.7) | Population-based randomized | US | Lung | 135 | Early + advanced | - | MVA | ✓ | - | - | 12 |
| Abdullah/2011<sup>22</sup> | 5036 | 567 (11) | Retrospective (cohort) | Various | Various | - | - | - | MVA | - | ✓ | - | 5 |
| Abrahamson/2006<sup>23</sup> | 1254 | - | Retrospective | US | Breast | - | Early + advanced | - | MVA | ✓ | - | - | 5 |
| Abukabar/2018<sup>24</sup> | 3012 | 433 (13) | Retrospective | Malaysia | Breast | 24 | Early + advanced | S ± RT ± CT | MVA | ✓ | - | ✓ | 6 |
| Ademuyiawa/2011<sup>25</sup> | 418 | 164 (39.2) | Retrospective | US | Breast | 37.2 | Early | S | MVA | ✓ | - | ✓ | 6 |
| Akinyemiju/2018<sup>26</sup> | 22514 | 8786 (39) | Prospective | US | Various | 78 | - | - | MVA | - | ✓ | - | 8 |
| Aldrich/2013<sup>28</sup> | 501 | 126 (25.1) | Prospective (cohort) | US | NSCLC | 16 | Early + advanced | - | MVA | ✓ | - | - | 7 |
| Alsaker/2011<sup>29</sup> | 2640 | 432 (16.3) | Retrospective | Norway | Breast | 69 | Early + advanced | - | MVA | - | ✓ | - | 7 |
| Arce-Salinas/2014<sup>30</sup> | 819 | 596 (74) | Retrospective | Mexico | Breast | 28 | Advanced | CT | UVA | ✓ | - | ✓ | 6 |
| Arem/2013<sup>31</sup> | 1400 | 610 (43) | Retrospective | US | Uterus | 61.2 | Early + advanced | - | MVA | ✓ | ✓ | - | 7 |
| Ata/2018<sup>32</sup> | 8352 | 2841 (34) | Retrospective | US | HCC | 60 | - | Liver transplantation | MVA | ✓ | ✓ | - | 7 |
| Bandera/2015<sup>33</sup> | 1846 | 547 (29.6) | Cohort study | US | Ovarian | - | Early + advanced | CT | MVA | ✓ | ✓ | - | 5 |
| Bassett/2012<sup>35</sup> | 16525 | 247 (18) | Prospective (cohort) | Australia | Prostate | 180 | - | - | MVA | - | ✓ | - | 12 |
| Beasley/2012<sup>27</sup> | 13302 | 2330 (17.5) | Metanalysis | US, China | Breast | - | Early | All | MVA | ✓ | ✓ | ✓ | - |
| Blair/2019<sup>36</sup> | 859 | 195 (22.7) | Cohort study | US | Breast | 94 | Early + advanced | CT, RT, HT | MVA | - | ✓ | - | 8 |
| Boggs/2011<sup>37</sup> | 51695 | 23656 (46) | Prospective | US | Various | - | - | - | MVA | - | ✓ | - | 8 |
| Bonn/2014<sup>38</sup> | 4376 | 483 (11) | Retrospective | Sweden | Prostate | 48 | Early | S, RT | MVA | ✓ | ✓ | ✓ | 6 |
| Boyle/2013<sup>39</sup> | 879 | 258 (29.4) | Retrospective | Australia | CRC | 67.2 | Early | - | MVA | ✓ | ✓ | ✓ | 7 |
| Braithwaite/2010<sup>40</sup> | 2202 | 500 (22.7) | Retrospective | US | Breast | 88 | Early | - | MVA | ✓ | ✓ | - | 8 |
| Campbell/2015<sup>44</sup> | 5615 | 1483 (26.4) | Prospective | Various | CRC | - | Early + advanced | S, CT | MVA | ✓ | ✓ | - | 5 |
| Caan/2008<sup>43</sup> | 1692 | 409 (24.2) | Retrospective | US | Breast | - | Early | S / adjuvant | MVA | ✓ | ✓ | ✓ | 5 |
| Carr/2018<sup>45</sup> | 521 | - | Retrospective | Italy | HCC | - | Early + advanced | - | MVA | ✓ | - | - | 5 |
| Cecchini/2016<sup>46</sup> | 5265 | 1794 (34) | Phase III (NSABP-B30) | US | Breast | 108 | Early | CT | MVA | ✓ | - | - | - |
| Cecchini/2016<sup>46</sup> | 2102 | 664 (31.6) | Phase III (NSABP-B31) | US | Breast | 99.6 | Early | CT + TTZ | MVA | ✓ | - | - | - |
| Cecchini/2016<sup>46</sup> | 3311 | 1186 (35.8) | Phase III (NSABP-B34) | US | Breast | 100.8 | Early | BPS vs Placebo* | MVA | ✓ | - | - | - |
| Cecchini/2016<sup>46</sup> | 4860 | 1917 (39.4) | Phase III (NSABP-B38) | US | Breast | 70.8 | Early | CT | MVA | ✓ | - | - | - |
| Céspedes Feliciano/2017<sup>47</sup> | 2470 | - | Retrospective | US | CRC | 72 | Early | - | MVA | ✓ | ✓ | - | 7 |
| Chang/2000<sup>48</sup> | 177 | 64 (36.2) | Retrospective | US | Breast | 100 | Advanced | Induction CT / S | MVA | ✓ | - | - | 8 |
| Chen/2010<sup>49</sup> | 5042 | 283 (5.6) | Retrospective | China | Breast | 6.5 | Early + advanced | S, CT, RT, HT | MVA | ✓ | ✓ | - | 6 |
| Hung/2018<sup>34</sup> | 33551 | 2362 (7.1) | Retrospective (cohort) | Taiwan | Solid cancers | 43.8 | Early + advanced | S | MVA | ✓ | ✓ | - | 6 |
| Chromecki/2012<sup>50</sup> | 4118 | 1739 (42.2) | Retrospective | Various | Bladder | 44 | Early | S ± adjuvant CT | MVA | ✓ | ✓ | ✓ | 7 |
| Chung/2017<sup>51</sup> | 8742 | 75 (8.5) | Retrospective | South Korea | Breast | 92 | Early | All | MVA | ✓ | ✓ | - | 9 |
| Clark/2013<sup>41</sup> | 99 | 40 (41) | Retrospective | US | CRC | 39.4 | Early | CT + RT | MVA | ✓ | - | ✓ | 6 |
| Cleveland/2012<sup>42</sup> | 1447 | 319 (22) | Prospective (case-control) | US | Breast | - | - | - | MVA | ✓ | ✓ | - | 8 |
| Connor/2016<sup>52</sup> | 2478 | 142 (5.7) | Prospective (registry) | US | Breast | 129.6 | Early + advanced | - | MVA | ✓ | ✓ | - | 12 |
| Conroy/2011<sup>53</sup> | 3842 | 901 (23) | Prospective (cohort) | US | Breast | 74.4 | Early + advanced | S ± CT/HT and/or RT | MVA | ✓ | ✓ | - | 11 |
| Copson/2014<sup>56</sup> | 2843 | 533 (18.7) | Prospective | UK | Breast | 70.4 | Early + advanced | S, RT, CT, HT | MVA | ✓ | - | ✓ | 11 |
| Cortellini/2019<sup>54</sup> | 976 | 377 (38.6) | Retrospective | Italy | Various | 17.2 | Advanced | Anti-PD-1/PD-L1 | MVA | ✓ | - | ✓ | 6 |
| Crozier 2013<sup>55</sup> | 3017 | 1298 (43) | Retrospective | US | Breast | 63.6 | Early | - | MVA | - | ✓ | ✓ | 8 |
| Dahdaleh/2018<sup>57</sup> | 1543 | 529 (34.3) | Retrospective (cohort) | US | CRC | 30.9 | Early | S, adjuvant CT | MVA | ✓ | - | ✓ | 6 |
| Dalal/2012<sup>59</sup> | 41 | 8 (20) | Prospective | US | Pancreas | - | Advanced | CTRT | MVA | ✓ | - | - | 7 |
| Dal Maso/2008<sup>58</sup> | 1453 | 172 (11.8) | Retrospective | Italy | Breast | - | Early | S / adjuvant | UVA | ✓ | ✓ | - | 5 |
| Dawood/2012<sup>60</sup> | 2311 | 825 (35.7) | Retrospective | US | Breast (TN) | 39 | Early | S | MVA | ✓ | - | ✓ | 6 |
| Dickerman/2017<sup>61</sup> | 5158 | 564 (10.9) | Retrospective | US | Prostate | - | Early | All | MVA | - | ✓ | - | 5 |
| Dignam/2003<sup>63</sup> | 3385 | 395 (11.67) | Retrospective | US | Breast | - | Early | S + HT | MVA | ✓ | ✓ | ✓ | 5 |
| Dignam/2006<sup>64</sup> | 4077 | 1056 (25.9) | Retrospective | US | Breast | - | - | - | MVA | ✓ | ✓ | ✓ | 5 |
| Dignam/2006<sup>65</sup> | 4288 | 812 (9.5) | Retrospective | North America | CRC | - | Early | CT | MVA | ✓ | ✓ | ✓ | 5 |
| Doria-Rose/2006<sup>66</sup> | 633 | 96 (15.2) | Retrospective | US | CRC (women) | 112.8 | Early | S / others | MVA | ✓ | ✓ | - | 8 |
| Drake/2017<sup>67</sup> | 7061 | 3220 (45.6) | Prospective (cohort) | Sweden | Various | 202 | Early + advanced | All | MVA | ✓ | ✓ | - | 12 |
| Efstathiou/2008<sup>62</sup> | 945 | 145 (15.3) | Prospective | US | Prostate | 97.2 | Advanced | RT +/- Goserelin | MVA | ✓ | ✓ | - | 11 |
| Emaus/2009<sup>69</sup> | 1364 | 147 (10.78) | Retrospective | Norway | Breast | 98.4 | Early + advanced | - | MVA | ✓ | ✓ | - | 8 |
| Farris/2018<sup>71</sup> | 987 | 192 (19.4) | Prospective (cohort) | Canada | Prostate | 228 | Early + advanced | S, RT, HT | MVA | ✓ | ✓ | - | 12 |
| Fedirko/2014<sup>72</sup> | 3924 | 689 (17.5) | Prospective | Western Europe | Colorectal | 49 | Early + advanced | - | MVA | ✓ | ✓ | - | 7 |
| Feliciano/2017<sup>73</sup> | 1559 | 471 (30.2) | Retrospective | US | Breast | 108 | Early | All | MVA | - | ✓ | ✓ | 8 |
| Ferro/2018<sup>74</sup> | 1155 | 224 (21.4) | Retrospective | Italy | Bladder | 48 | Early | TURBT + BCG | MVA | - | ✓ | ✓ | 7 |
| Froehner/2014<sup>75</sup> | 2131 | 356 (16.7) | Retrospective | Germany | Prostate | 110 | Early | All | UVA, MVA | ✓ | ✓ | - | 9 |
| Frumovitz/2014<sup>76</sup> | 3086 | 1026 (33.2) | Retrospective | US | Cervical | 133 | Early + advanced | S, RT | MVA | ✓ | ✓ | - | 9 |
| Gama/2017<sup>121</sup> | 1279 | 243 (21) | Retrospective | Canada | H&N | 30 | Early + advanced | RT, S, CT | MVA | ✓ | - | ✓ | 6 |
| Genkinger/2015<sup>77</sup> | 1096492 | - | Cohort study | US | Pancreas | 152.4 | - | - | MVA | - | ✓ | - | 11 |
| Gong/2007<sup>78</sup> | 752 | 128 (17) | Retrospective | US | Prostate | 116.4 | Early + advanced | S / ADT / RT / others | MVA | - | ✓ | - | 9 |
| Gong/2012<sup>70</sup> | 510 | 51 (10) | Retrospective | US | Pancreas | 121.2 | Early + advanced | All | MVA | ✓ | - | - | 9 |
| Goodwin/2012<sup>79</sup> | 535 | - | Retrospective | Canada | Breast | 145.2 | Early | S ± adjuvant CT and/or HT | MVA | ✓ | - | ✓ | 9 |
| Grossberg/2016<sup>80</sup> | 190 | 65 (34.2) | Retrospective | US | H&N | 68.6 | Early | CT + RT | MVA | ✓ | ✓ | ✓ | 7 |
| Halabi/2007<sup>82</sup> | 1296 | 405 (31.2) | Retrospective | US | Prostate (androgen-independent) | 33.8 | Early + advanced | - | MVA | ✓ | ✓ | - | 6 |
| Han/2010<sup>84</sup> | 2511 | 211 (8.4) | Retrospective | US | Prostate | 156 | Early | All | UVA | ✓ | - | - | 9 |
| Han/2014<sup>85</sup> | 13901 | 708 (5.09) | Retrospective | US | Various | - | - | - | MVA | - | ✓ | - | 5 |
| Xu /2018<sup>215</sup> | 644 | 92 (14.3) | Retrospective | China | Upper tract urothelial | 39 | Early | S | MVA | ✓ | ✓ | ✓ | 6 |
| He/2012<sup>86</sup> | 1983 | 546 (27.5) | Retrospective | US | Breast | 47.6 | Early + advanced | All | MVA | ✓ | - | - | 7 |
| Hellmann/2010<sup>87</sup> | 528 | 76 (14.4) | Prospective | Denmark | Breast | 93.6 | Early + advanced | - | MVA | ✓ | ✓ | - | 10 |
| His/2016<sup>88</sup> | 3160 | 194 (6.1) | Prospective | France | Breast | 109.2 | Early | - | MVA | ✓ | ✓ | ✓ | 11 |
| Ho/2012<sup>89</sup> | 1038 | 337 (32.4) | Retrospective | US | Prostate | 41 | Early + advanced | S | MVA | - | - | ✓ | 6 |
| Houdek/2019<sup>90</sup> | 261 | 71 (8.6) | Retrospective | - | STS | 48 | - | RT / none | MVA | ✓ | - | ✓ | 6 |
| Iyengar/2014<sup>94</sup> | 155 | 30 (19) | Retrospective | US | Tongue | - | Early | S | MVA | ✓ | ✓ | ✓ | 5 |
| Izumida/2019<sup>95</sup> | 10824 | 235 (2) | Cohort study | China | Various | 220.8 | - | - | MVA | - | ✓ | - | 12 |
| Janssen/2015<sup>96</sup> | 927 | - | Retrospective | US | Various | - | Early + advanced | - | MVA | ✓ | - | - | 5 |
| Jayasekara/2018<sup>97</sup> | 724 | 164 (22.7) | Cohort study | Australia | CRC | 108 | Early | - | MVA | ✓ | ✓ | - | 11 |
| Jenkins/2018<sup>98</sup> | 502631 | 12539 (24.9) | Cohort study | UK | Various | 93.6 | - | - | MVA | ✓ | - | - | 7 |
| Jeon/2015<sup>99</sup> | 41021 | 1632 (4) | Prospective (registry) | Korea | Breast | 92 | Early | CT, HT | MVA | ✓ | ✓ | - | 11 |
| Jiralerspong/2013<sup>100</sup> | 6342 | 1779 (30) | Retrospective | US | Breast | 64.8 | Early | - | MVA | ✓ | ✓ | ✓ | 7 |
| Kaidar-Person/2015<sup>101</sup> | 184 | 46 (25) | Retrospective | Israel | CRC | 27.6 | Advanced | CT, Bevacizumab | MVA | ✓ | - | ✓ | 6 |
| Kalb/2019<sup>102</sup> | 612 | 127 (20.7) | Retrospective | Germany | CRC | 58 | Early | CT, RT, S | MVA | ✓ | - | ✓ | 7 |
| Katzmarzyk/2012<sup>103</sup> | 10522 | 1972 (19) | Retrospective | Canada | Various | 168 | Early + advanced | All | MVA | - | ✓ | - | 8 |
| Kawai/2016<sup>104</sup> | 20090 | 897 (4.5) | Prospective (registry) | Japan | Breast | 80.4 | Early | CT, HT | MVA | ✓ | ✓ | ✓ | 10 |
| Keegan/2010<sup>105</sup> | 4153 | 127 (3) | Retrospective | US | Breast | - | - | - | MVA | ✓ | - | - | 5 |
| Keizman/2014<sup>106</sup> | 278 | 67 (24.1) | Retrospective | Israel | RCC | 55 | Advanced | TKI | MVA | ✓ | - | ✓ | 7 |
| Kelly/2017<sup>107</sup> | 7822 | 1612 (20.6) | Retrospective | US | Prostate | 156 | Early + advanced | All | MVA | - | ✓ | - | 9 |
| Kenfield/2016<sup>108</sup> | 112185 | 9984 (8.9) | Prospective | US | Prostate | 170 | - | - | MVA | - | ✓ | - | 12 |
| Khan/2017<sup>109</sup> | 822 | - | Retrospective | US | Prostate | 60 | Early + advanced | All | MVA | - | - | ✓ | 7 |
| Kichenadasse/2019<sup>110</sup> | 1434 | 239 (7) | Prospective | Various | NSCLC | - | Advanced | Atezolizumab vs Docetaxel | UVA | ✓ | - | ✓ | 7 |
| Kitahara/2014<sup>111</sup> | 313575 | 9564 (3.04) | Retrospective | Various | Various | - | - | - | MVA | - | ✓ | - | 5 |
| Kotsopoulos/2012<sup>113</sup> | 1423 | 230 (17.5) | Retrospective | Canada | Ovarian | 120 | Early + advanced | All | MVA | - | ✓ | - | 9 |
| Kristensen/2017<sup>114</sup> | 4330 | - | Retrospective | Denmark | Endometrial | - | Early + advanced | S | MVA | ✓ | - | - | 5 |
| Kwan/2012<sup>115</sup> | 14948 | 2440 (16.3) | Prospective (cohort) | US | Breast | 93.6 | Early | S ± adjuvant CT and/or HT and/or RT | MVA | ✓ | ✓ | - | 11 |
| Kwan/2013<sup>116</sup> | 11351 | 3405 (30) | 3 pooled case-control, 3 prospective cohort | US | Breast | 132 | Early + advanced | All | MVA | ✓ | ✓ | - | - |
| Ladoire/2014<sup>117</sup> | 5009 | 666 (13.3) | Pooled analysis (2 phase III) | France | Breast | 70.8 | Early | CT | MVA | ✓ | - | ✓ | - |
| Larsen/2015<sup>118</sup> | 1229 | 167 (14) | Prospective (cohort) | Denmark | Breast | 115.2 | Early | - | MVA | ✓ | - | - | 12 |
| Lee/2010<sup>120</sup> | 2750 | 120 (4.3) | Retrospective | South Korea | RCC | 34.8 | Early | S | MVA | - | ✓ | ✓ | 6 |
| Leung/2011<sup>122</sup> | 58931 | 3520 (5.97) | Prospective | Japan | Lung | - | - | - | MVA | - | ✓ | - | 7 |
| Li/2009<sup>123</sup> | 841 | 163 (19.38) | Retrospective | US | Pancreas | - | Early + advanced | - | MVA | ✓ | - | - | 5 |
| Lin/2013<sup>124</sup> | 799542 | 19988 (2.5) | Retrospective | Various | Pancreas | 37.2 | - | - | MVA | - | ✓ | - | 6 |
| Loi/2005<sup>125</sup> | 1101 | 131 (11.9) | Retrospective | Australia | Breast | 60 | Early | S / adjuvant | MVA | ✓ | - | ✓ | 6 |
| Ma/2008<sup>126</sup> | 2546 | 87 (3.4) | Retrospective | US | Prostate | 84 | Early + advanced | - | MVA | ✓ | ✓ | - | 7 |
| Maj-Hes/2017<sup>127</sup> | 6519 | 2462 (37.7) | Retrospective | Austria | Prostate | 28 | Early | S | MVA | - | - | ✓ | 6 |
| Martini/2020<sup>128</sup> | 90 | 23 (25.5) | Retrospective | US | Various | - | Advanced | IT | MVA | ✓ | - | ✓ | 5 |
| Maskarinec/2011<sup>129</sup> | 382 | 71 (18.5) | Prospective (cohort) | Hawaii | Breast | 158.4 | Early + advanced | S ± adjuvant CT and/or HT | MVA | ✓ | ✓ | - | 12 |
| Mathur/2010<sup>130</sup> | 279 | 97 (35) | Retrospective | US | Various | 31 | Advanced | Hepatectomy | MVA | ✓ | - | ✓ | 6 |
| Matsuo/2016<sup>131</sup> | 665 | 459 (69) | Retrospective | US | Uterus | 36.4 | Early + advanced | S + CTRT | MVA | ✓ | - | ✓ | 6 |
| Mazzarella/2013<sup>133</sup> | 1250 | 101 (8.1) | Retrospective | Europe | Breast | - | Early | S, RT, CT | MVA | ✓ | - | ✓ | 5 |
| McCullough/2005<sup>134</sup> | 430236 | 5433 (1.3) | Prospective | US | Breast | 240 | Early + advanced | All | MVA | - | ✓ | - | 12 |
| McCullough/2016<sup>135</sup> | 1308 | 301 (23) | Population-based | US | Breast | 180 | Early | - | MVA | ✓ | ✓ | - | 12 |
| McMahon/2017<sup>83</sup> | 1080 | - | Retrospective | US | Liver | 123.6 | Early + advanced | All | MVA | - | ✓ | - | 9 |
| McQuade/2019<sup>136</sup> | 1918 | 513 (26.7) | Pooled analysis | US | Melanoma | - | Advanced | CT, IT, TT | MVA | ✓ | - | ✓ | - |
| Melhem-Bertrandt/2011<sup>138</sup> | 1413 | 460 (33) | Retrospective | US | Breast (TN) | 59 | Early | S ± CT | MVA | ✓ | - | ✓ | 7 |
| Meyer/2015<sup>139</sup> | 35703 | 2,820 (7.9) | Population-based | Switzerland | Various | - | Early + advanced | - | MVA | ✓ | - | - | 7 |
| Meyerhardt/2003<sup>140</sup> | 3561 | 500 (16.8) | Cohort study | US | CRC | 112 | Early | S + CT | MVA | ✓ | - | ✓ | 12 |
| Meyerhardt/2004<sup>141</sup> | 1688 | 306 (18) | Cohort study | US | CRC | 118 | Early | S + CT | MVA | ✓ | - | ✓ | 12 |
| Meyerhardt/2008<sup>142</sup> | 1043 | 236 (22.6) | Prospective | US and Canada | CRC | 63.6 | Early | S / adjuvant | MVA | ✓ | - | ✓ | 8 |
| Minlikeeva/2019<sup>112</sup> | 7022 | 1557 (22.2) | Retrospective (pool data) | US and Australia | Ovarian | - | Early + advanced | - | MVA | ✓ | - | ✓ | 5 |
| Moller/2014<sup>143</sup> | 26877 | 4140 (15) | Cohort study | UK | Prostate | 43.2 | Early + advanced | - | MVA | - | ✓ | - | 11 |
| Montgomery/2007<sup>144</sup> | 1006 | 160 (16) | Retrospective | US | Prostate (androgen-dependent) | - | Advanced | Bilateral orchiectomy ± Flutamide | MVA | ✓ | - | ✓ | 5 |
| Montgomery/2007<sup>144</sup> | 671 | 253 (38) | Retrospective | US | Prostate (androgen-independent) | - | Advanced | Mitoxantrone/Prednisone vs Docetaxel/Estramustine | MVA | ✓ | - | ✓ | 5 |
| Morikawa/2011<sup>145</sup> | 1060 | 200 (19) | Prospective (cohort) | US | CRC | 162 | Early + advanced | All | MVA | ✓ | ✓ | - | 12 |
| Nagle/2018<sup>148</sup> | 1359 | 568 (41.7) | Retrospective | Australia | Endometrial | 85.2 | Early + advanced | S ± CT | MVA | ✓ | ✓ | - | 8 |
| Nakagawa/2016<sup>149</sup> | 1311 | 25 (1.9) | Retrospective | Japan | NSCLC | 59 | Early | S | MVA | ✓ | - | ✓ | 7 |
| Nechuta/2010<sup>150</sup> | 71243 | 8264 (11.6) | Cohort study | UK | Various | 109.2 | - | - | MVA | - | ✓ | - | 12 |
| Nicholas/2014<sup>151</sup> | 490 | 203 (41.4) | Retrospective | US | Endometrial | 54 | Early + advanced | S | MVA | ✓ | - | - | 6 |
| Nichols/2009<sup>152</sup> | 3993 | 639 (16) | Retrospective | US | Breast | 76.8 | Early | S | MVA | ✓ | ✓ | - | 7 |
| Nonemaker/2009<sup>153</sup> | 2054 | 50 (12.5) / 46 (8.4) | Retrospective | Africa / Europe and America | Lung | - | - | - | MVA | ✓ | ✓ | - | 5 |
| Nur/2019<sup>154</sup> | 49259 | 202 (4.1) | Retrospective | Sweden | Breast | 91.6 | Early | S | MVA | ✓ | ✓ | - | 8 |
| Ogino/2009<sup>155</sup> | 546 | 84 (16) | Retrospective | US | CRC | - | Early + advanced | - | MVA | ✓ | ✓ | - | 5 |
| Oh/2011<sup>156</sup> | 747 | 251 (33.7) | Cohort study | Korea | Breast | 62.2 | Early | S ± CT | MVA | - | - | ✓ | 10 |
| Olson/2010<sup>157</sup> | 475 | 108 (23) | Retrospective | US | Pancreas | - | Early + advanced | S | MVA | ✓ | ✓ | - | 5 |
| Oudanonh/2019<sup>158</sup> | 3747 | 1790 (47.8) | Retrospective (registry) | Canada | Breast | - | Early | HT, CT, RT, antiHER2 | MVA | ✓ | - | - | 5 |
| Pajares/2013<sup>159</sup> | 5683 | 1376 (24.4) | Retrospective | Spain | Breast | - | Early | CT, HT, S | MVA | ✓ | ✓ | ✓ | 5 |
| Parker/2006<sup>160</sup> | 970 | 336 (34.6) | Retrospective | US | RCC | 56.4 | Early | S | MVA | ✓ | ✓ | - | 7 |
| Parr 2010<sup>161</sup> | 401215 | 16978 (4.23) | Retrospective | All | Various | - | - | - | MVA | ✓ | - | - | 5 |
| Patel/2015<sup>162</sup> | 1174 | 462 (39.4) | Retrospective | Australia | CRC | - | Advanced | CT | MVA | ✓ | - | - | 5 |
| Pelser/2014<sup>163</sup> | 5727 | - | Retrospective | US | CRC | - | Early + advanced | S, RT, CT | MVA | ✓ | ✓ | - | 5 |
| Pfeiler/2013<sup>164</sup> | 1509 | 315 (21) | Retrospective | Austria | Breast | 60 | Early | - | MVA | ✓ | ✓ | ✓ | 7 |
| Pierce/2007<sup>166</sup> | 1490 | 380 (25.5) | Prospective | US | Breast | 80.4 | Early | S / adjuvant | MVA | ✓ | - | - | 8 |
| Potharaju/2018<sup>167</sup> | 392 | 40 (10.2) | Retrospective | India | GBM | 48.6 | - | S + RT + TMZ | MVA | ✓ | - | - | 6 |
| Previs/2014<sup>168</sup> | 81 | 28 (34) | Retrospective | US | Ovarian | - | Early + advanced | S, RT | MVA | ✓ | ✓ | ✓ | 5 |
| Prizment/2010<sup>169</sup> | 1096 | 295 (26.9) | Retrospective (registry) | US | CRC | 240 | Early + advanced | CT, RT, S | MVA | ✓ | ✓ | - | 9 |
| Probst-Hensch/2010<sup>170</sup> | 855 | 72 (19.8) | Retrospective | Switzerland | Breast | 43.8 | Early | S ± HT | MVA | ✓ | - | - | 6 |
| Psutka/2015<sup>171</sup> | 387 | 166 (43) | Retrospective | US | RCC | 86.4 | Early | S | MVA | ✓ | ✓ | ✓ | 8 |
| Qi/2009<sup>172</sup> | 420 | 79 (22.7) | Retrospective | US | Lung | - | Advanced | All | MVA | ✓ | - | - | 5 |
| Qinchao Hu/2019<sup>91</sup> | 576 | 33 (5.7) | Retrospective | China | H&N | 64 | Early | S | MVA | - | ✓ | ✓ | 7 |
| Roque/2016<sup>173</sup> | 128 | 72 (56.3) | Retrospective | US | Leiomyosarcoma | 49 | Early + advanced | All | MVA | ✓ | - | ✓ | 6 |
| Rosenberg/2010<sup>92</sup> | 2640 | 376 (14.24) | Retrospective | Sweden | Breast | - | Early + advanced | S, CT, HT, RT | MVA | - | ✓ | - | 5 |
| Rudman/2016<sup>174</sup> | 273 | 59 (21.6) | Retrospective | UK | Prostate | 139.2 | Early | HT | MVA | ✓ | ✓ | - | 9 |
| Ruterbusch/2014<sup>93</sup> | 627 | 184 (29) | Retrospective | US | Uterus | - | Early + advanced | S ± CT | MVA | ✓ | ✓ | - | 5 |
| Sanchez/2019<sup>175</sup> | 730 | 249 (34) | Cohort | US | RCC | - | Early + advanced | Systemic therapy + S | MVA | ✓ | - | - | 6 |
| Sasazuki/2011<sup>176</sup> | 353422 | 7327 (2) | Prospective (cohort) | Japan | Various | 150 | - | - | MVA | - | ✓ | - | 12 |
| Schiffmann/2017<sup>177</sup> | 16014 | 2403 (15) | Retrospective | Germany | Prostate | 36.4 | Early | S | MVA | - | - | ✓ | 6 |
| Schlesinger/2014<sup>178</sup> | 2143 | 397 (19) | Prospective | Germany | CRC | 42 | Early + advanced | - | MVA | ✓ | - | - | 6 |
| Seidelin/2016<sup>179</sup> | 3638 | 984 (27) | Population-based | Denmark | Uterus | - | Early + advanced | - | MVA | ✓ | - | - | 7 |
| Senie/1992<sup>180</sup> | 923 | 207 (22) | Prospective | US | Breast | 120 | Early | - | MVA | - | - | ✓ | 12 |
| Shah/2015<sup>181</sup> | 242 | 59 (24.5) | Prospective | US | CRC | - | Advanced | S, CT | MVA | ✓ | - | - | 7 |
| Shepshelovich/2019<sup>183</sup> | 29217 | 418 (1.4) | Pooled analysis | Canada | Lung | - | Early + advanced | - | MVA | ✓ | - | - | - |
| Siegel/2013<sup>119</sup> | 853 | 216 (25) | Prospective | US | Brain | 19 | Early | - | MVA | ✓ | - | - | 8 |
| Silventoinen/2014<sup>184</sup> | 734438 | 9187 (1.2) | Retrospective | Finland, Sweden | Various | 403.2 | - | - | MVA | ✓ | ✓ | - | 9 |
| Sinicrope/2012<sup>185</sup> | 2693 | 630 (23) | Randomized trial | US | CRC | - | Early | S ± CT | MVA | ✓ | - | ✓ | 5 |
| Sinicrope/2013<sup>81</sup> | 25291 | 4463 (17.6) | Retrospective | US | CRC | 93.6 | Early | - | MVA | ✓ | - | ✓ | 8 |
| Song/2012<sup>132</sup> | 135745 | - | Prospective | Europe | Various | 201.6 | - | - | MVA | - | ✓ | - | 12 |
| Sorbye/2012<sup>137</sup> | 342 | 67 (19.5) | Retrospective | Europe | CRC | - | Advanced | S ± CT | MVA | - | - | ✓ | - |
| Spangler/2007<sup>186</sup> | 924 | 286 (31) | Prospective | Various | Prostate | 36 | Early | S | MVA | - | - | ✓ | 10 |
| Sparano/2012<sup>187</sup> | 4817 | 1745 (46) | Retrospective (phase III) | US | Breast | 95 | Early | S + CT + HT | MVA | ✓ | ✓ | ✓ | - |
| Sparano/2012<sup>188</sup> | 6885 | 2547 (37) | Retrospective (3 phase III) | US | Breast | 95 | Early | S ± CT ± HT | MVA | ✓ | ✓ | ✓ | 9 |
| Spiess/2012<sup>189</sup> | 99 | 43 (43) | Retrospective | US | RCC | 44.4 | Early + advanced | S | MVA | ✓ | - | - | 6 |
| Spreafico/2017<sup>190</sup> | 564 | 76 (13) | Retrospective | Canada | Esophagus | 32.5 | Early + advanced | All | MVA | ✓ | - | ✓ | 6 |
| Su/2013<sup>191</sup> | 1030 | 312 (30.3) | Randomized study | US | Breast | - | Early | S ± CT ± HT | MVA | - | - | ✓ | - |
| Sun/2015<sup>192</sup> | 1109 | 410 (37) | Population-based | US | Breast | 162 | Early | - | MVA | ✓ | ✓ | - | 8 |
| Sun/2018<sup>193</sup> | 1017 | 192 (18.9) | Retrospective | China | Breast | 80 | Early | CT, HT, RT, S | MVA | ✓ | - | ✓ | 8 |
| Sundelof/2009<sup>194</sup> | 580 | 55 (9.5) | Retrospective | Sweden | Esophagus | - | Early + advanced | - | MVA | ✓ | - | - | 5 |
| Taghizadeh/2015<sup>195</sup> | 8645 | 683 (7.9) | Cohort study | Netherlands | Various | 480 | - | - | MVA | ✓ | - | - | 12 |
| Tait/2014<sup>196</sup> | 501 | 202 (40.3) | Retrospective | US | Breast | 40.1 | Early + advanced | S, CT | MVA | ✓ | - | ✓ | 6 |
| Thrift/2013<sup>197</sup> | 783 | 263 (33) | Retrospective | Australia | Esophagus / Gastric | 76.8 | Early + advanced | S ± CT/RT/BSC | MVA | ✓ | - | - | 8 |
| Todo/2014<sup>198</sup> | 716 | 99 (13.8) | Retrospective | Japan | Endometrial | 74 | Early + advanced | S, CT | MVA | ✓ | ✓ | - | 7 |
| Trivers/2005<sup>200</sup> | 1142 | 156 (4.2) | Retrospective | US | Esophagus / Gastric | - | Early + advanced | S / others | MVA | ✓ | - | ✓ | 5 |
| Tsai/2010<sup>201</sup> | 795 | 103 (13) | Retrospective | US | Pancreatic | - | Early + advanced | S | MVA | ✓ | - | - | 5 |
| Tseng/2013<sup>146</sup> | 89056 | - | Prospective | Taiwan | Various | 144 | Early + advanced | All | MVA | - | ✓ | - | 12 |
| Tseng/2016<sup>202</sup> | 92546 | - | Retrospective | Taiwan | Various | 204 | Early + advanced | All | MVA | - | ✓ | - | 9 |
| Turner/2011<sup>203</sup> | 188699 | 22054 (12.4) | Prospective | US | Lung | 312 | - | - | MVA | - | ✓ | - | 12 |
| Tyler/2012<sup>204</sup> | 425 | 28 (6.6) | Prospective (case-control) | US | Ovarian | 116.4 | Early + advanced | All | MVA | ✓ | ✓ | - | 12 |
| Valentijn/2013<sup>205</sup> | 10247 | 1851 (18) | Retrospective | The Netherlands | Various | 64.8 | - | - | MVA | - | ✓ | - | 7 |
| Vidal/2017<sup>206</sup> | 4268 | 1372 (32.1) | Retrospective | US | Prostate | 81.6 | Early | S | MVA | - | ✓ | ✓ | 8 |
| Wang/2017<sup>208</sup> | 1452 | - | Retrospective | China | CRC | 40.8 | Early + advanced | All | MVA | - | ✓ | - | 6 |
| Wang/2019<sup>209</sup> | 250 | 81 (12) | Retrospective | US | Various | - | Advanced | IT | MVA | ✓ | - | ✓ | 5 |
| Warren/2016<sup>210</sup> | 878 | - | Retrospective | US | Breast | 129.6 | Early | S | UVA | - | - | ✓ | 9 |
| Widschwendter/2015<sup>211</sup> | 3754 | 788 (21) | Phase III (SUCCESS A) | Germany | Breast | - | Early | CT + HT | MVA | ✓ | - | ✓ | - |
| Wu/2015<sup>212</sup> | 333 | 118 (35.4) | Retrospective | US | Prostate | - | Advanced | CT | MVA | ✓ | - | - | 5 |
| Xiao/2014<sup>213</sup> | 5785 | 1680 (29) | Retrospective | China | Breast | 70 | Early | CT, HT | MVA | ✓ | - | - | 7 |
| Xie/2017<sup>147</sup> | 624 | - | Retrospective | China | Lung | 63.2 | Early | S | MVA | ✓ | - | - | 6 |
| Xu/2018<sup>214</sup> | 6197 | 1885 (30.4) | Cohort study | US | Various | 204 | - | - | MVA | - | ✓ | - | 12 |
| Yang/2008<sup>216</sup> | 635 | 81 (12.8) | Prospective | Europe | Ovarian | 96 | Early + advanced | - | MVA | - | ✓ | - | 10 |
| Yang/2019<sup>217</sup> | 2442 | 86 (3.5) | Retrospective | US | HCC | 50.5 | Early | S | MVA | ✓ | - | ✓ | 8 |
| Yano/2013<sup>218</sup> | 3641 | 792 (21.7) | Prospective | Japan | Various | 122 | - | - | MVA | - | ✓ | - | 12 |
| Yoon/2011<sup>219</sup> | 778 | 46 (19) | Retrospective | US | Esophagus (adeno) | 154.8 | Early | S ± adjuvant CT ± RT ± CTRT | MVA | ✓ | ✓ | ✓ | 9 |
| Yoon/2015<sup>220</sup> | 2987 | 417 (13.9) | Retrospective (cohort) | US | Endometrial | - | Early + advanced | S, CT, RT | MVA | ✓ | - | - | 5 |
| You/2015<sup>221</sup> | 1314 | - | Prospective (cohort) | China | Various | 52.7 | Early + advanced | - | MVA | ✓ | - | ✓ | 10 |
| Yu/1991<sup>165</sup> | 360 | 44 (12) | Retrospective | US | RCC | 53 | Early | S | MVA | ✓ | - | - | 7 |
| Yuan/2013<sup>222</sup> | 902 | 136 (15) | Prospective (cohort) | US | Pancreas | 480 | Early + advanced | - | MVA | ✓ | - | - | 12 |
| Zheng/2015<sup>223</sup> | 226 | 52 (23) | Cohort study | China | CRC | - | Early + advanced | - | MVA | ✓ | - | ✓ | 5 |

\*Clinical Trial Randomizing Breast Cancer Patients To Receive BFS Vs Placebo; Patients Received Concomitant Systemic Anticancer Treatment According To Physicians' Decision (Following Institutional Guidelines).
OS, Overall Survival ; CSS, Cancer Specific Survival; DFS, Disease-Free Survival; PFS, Progression-Free Survival; DDFI, Distant Disease Free Interval; US, United States Of America; UK, United Kingdom; NSCLC, Non-Small Cell Lung Cancer; HCC, Hepatocellular Carcinoma; RCC, Renal Cell Carcinoma; CRC, Colorectal Cancer; H&N, Head and Neck Tumors; STS, Soft Tissue Sarcoma; SCC, Squamous Cell Carcinoma; GBM, Glioblastoma; TN, Triple Negative; HR, Hormone Receptor; MVA, Multivariate Analysis; UVA, Univariate Analysis; S, Surgery; CT, Chemotherapy; HT, Hormone Therapy; IT, Immunotherapy; TKI, Tyrosine Kinase Inhibitor; TT, Targeted Therapy; TTZ, Trastuzumab; PD-1, Programmed Cell Death 1; PD-L1, Programmed Cell Death Ligand 1; ADT, Androgen Deprivation Therapy; BPS, Bisphosphonate; TMZ, Temozolomide; BCG, Bacillus Calmette-Guérin; RT, Radiotherapy; TURBT, Transurethral Resection of Bladder Tumor; BSC, Best Supportive Care

### Overall survival and obesity in cancer patients

N=170 studies reported data on OS. Because the heterogeneity test showed a high level of heterogeneity (I^2^ =79.7%, P<.01) among studies, a random-effects model was used for the analysis. OS of obese patients was significantly worse (HR =1.14, 95% CI: 1.09-1.19; P<01; Figure 2) compared to those of not obese patients.

**Figure 2.**
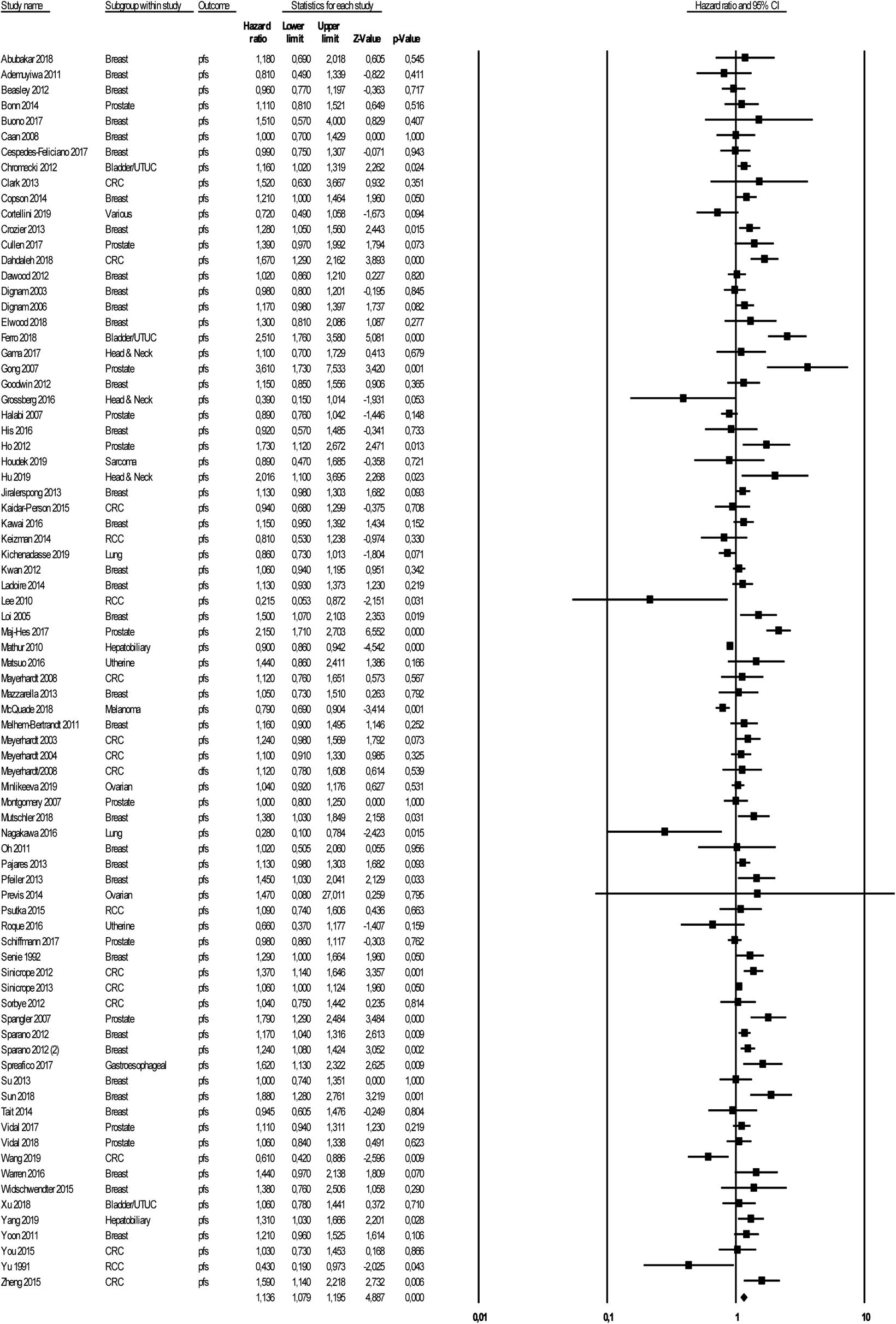
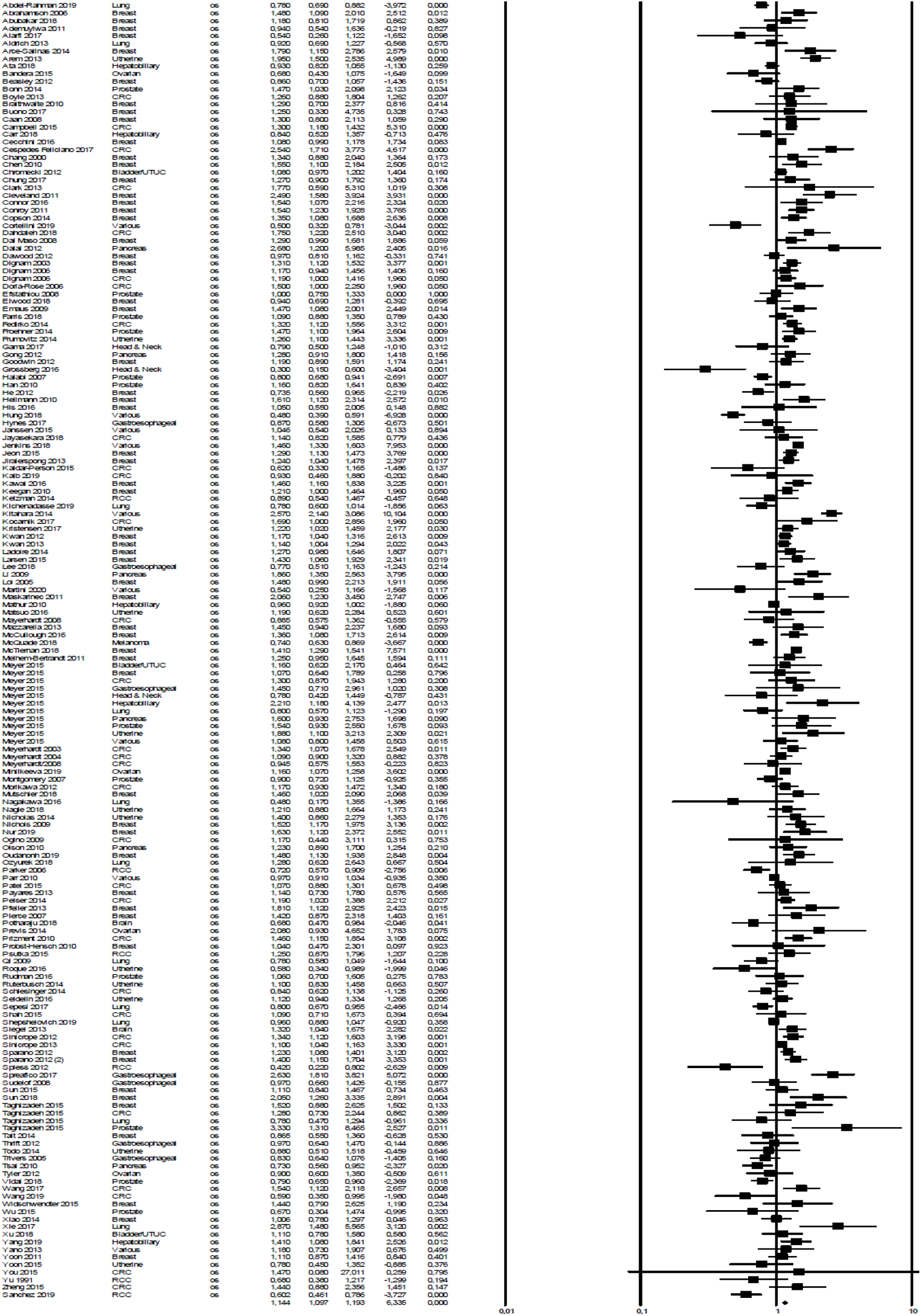
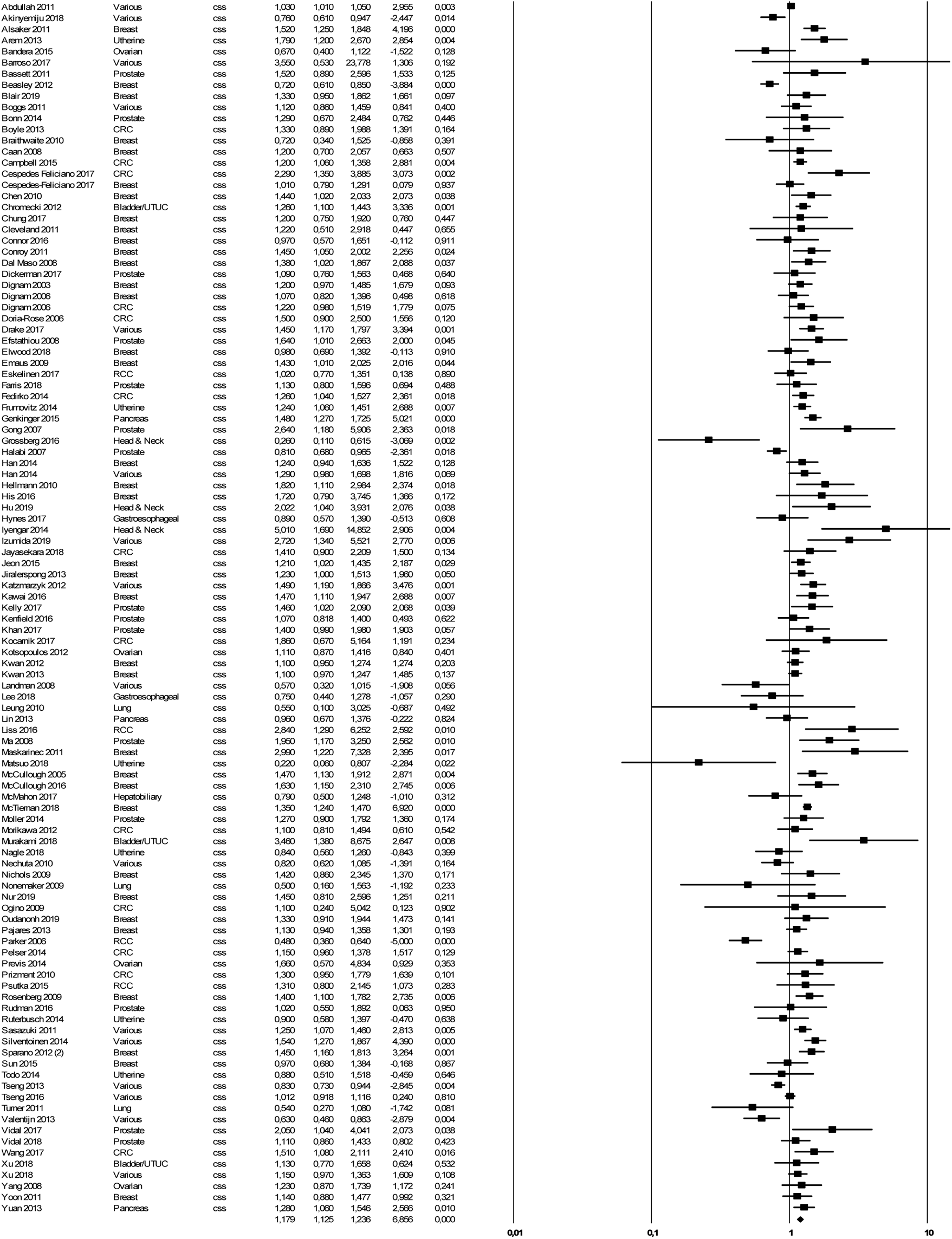

### Cancer-specific survival and obesity in cancer patients

Similarly, obesity reduced CSS in n=109 studies (HR=1.17, 95%CI 1.12-1.23; P<.01; Figure 3), which reported data on cancer mortality. Heterogeneity was high (I^2^=73.9%), so a random effect model was used

### Progression-free survival and obesity in cancer patients

Obese patients with cancer (n=79 studies) have a worse DFS/PFS than non-obese subjects (HR=1.13, 95%CI 1.07-1.19; P<0.01). Heterogeneity was high (I^2^=73.7%), so a random effect model was used.

### Subgroup analysis

A subgroup analysis for OS was performed according to type of disease (Suppl. file 1), type of study (retrospective vs prospective: HR=1.07, 95%CI 1.07-1.18, P<.01 vs HR=1.14, 95%CI 1.05-1.23, P<.01), duration of follow up (> vs < 10 years HR=1.16, 95%CI 0.86-1.58, P=.08 vs HR=1.23, 95%CI 0.84–3; P=.09), race (Non asiatic HR= 1.22, 95%CI 0.86-1.6, P=.06 vs HR=1.22, 0.74-1.72, P=.09) and stage of disease (early vs advanced: HR=1.20, 95%CI 0.99-1.25, P=.072 vs HR=1.2, 95% 1.12-1.28, P=.01).

### Publication bias

A funnel plot was used to assess publication bias in the studies evaluating OS in obese versus not obese cancer patients. No publication bias was detected (Begg’s test P=.23). Egger’s test was instead significant (P=0.03) (Suppl. file 2). According to the Trim and Fill method n=18 studies were placed to the left of the mean, and according the random effect model the final result was similar (HR=1.08, 95%CI 1.03-1.13). After the one study removed procedure the HR for ranged from 1.14 to 1.15.

## Discussion

This meta-analysis of published (observational and retrospective) studies shows that overall mortality is increased in obese patients with breast, colorectal and uterine cancers. Cancer mortality is increased in breast, colorectal, prostate and pancreatic cancers. Finally, relapse rate is increased in breast, colorectal, prostate and gastroesophageal cancers. The “obesity paradox”, that means the apparent better cancer mortality in obese patients, is observed in lung cancer and in melanoma, despite these data derive from n=12 studies only.

Various factors are potentially linked to increased cancer mortality in some malignancies. Hormonal factors, reduced physical activity, more lethal/aggressive disease behavior, metabolic syndrome and a potential undertreatment in obese subjects are possible reasons. It is well known that postmenopausal women with higher BMI have an increased breast cancer risk due to higher estrogen levels resulting from the peripheral conversion of estrogen precursors (from adipose tissue) to estrogen^224^. In these patients weight loss and exercise may reduce cancer risk through lowering exposure to breast cancer biomarkers^225^. In colorectal cancer, BMI before diagnosis was associated with increased all-cause, cardiovascular and colorectal cancer specific mortality^226^. The reason of this association is presently not completely understood although insulin, insulin-like growth factors, their binding proteins, chronic inflammation, oxidative stress, and impaired immune surveillance have been supposed to be causative factors^227^. In pancreatic cancer higher prediagnostic BMI was associated with more advanced stage at diagnosis, with 72.5% of obese patients presenting with metastatic disease versus 59.4% of healthy-weight patients (P = .02) in 2 large prospective cohort studies^222^. Lastly, in prostate cancer obesity may a consequence of androgen deprivation therapy but seems also related with more aggressive disease (i.e., Gleason score ≥7)^228^ or more advanced disease at diagnosis^229^.

Our results showed that obese lung cancer patients had significantly prolonged cancer specific survival and overall survival compared to non-obese patients. When considering these findings, we must take into account that 9 out of 11 evaluated studies included advanced/metastatic patients. Cancer cachexia mechanisms are not completely defined yet, but several evidences showed that the systemic inflammatory status induced either by the tumor and host-response is a key moment in its developing^230^. Lung cancers are indeed known to be aggressive diseases, and advanced patients are usually accompanied by poorer performance status and significant weight loss at the time of diagnosis, which underlies a systemic inflammatory response^231^. According to this view, a post-hoc analysis from three international phase III studies including advanced non-small cell lung cancer patients who received a platinum-based, first-line chemotherapy, revealed that weight gain during treatment was significantly associated with better clinical outcomes^232^. Intriguingly, a recent study showed that a decrease in BMI at lung cancer diagnosis (from the early adulthood) is a consistent marker of poor survival^183^. Considering these observations, an increased BMI might be considered a protective feature and potentially a marker of functional reserve for lung cancer patients. Recent evidences suggest a positive predictive role of an increased BMI for cancer patients receiving immune checkpoint inhibitors across different malignancies, including lung cancer^54^. Interestingly, the only study with lung cancer patients who received an immune checkpoint inhibitor (atezolizumab), revealed that the OS benefit for obese patients was significantly higher within the atezolizumab-treated cohort, compared to the doxetaxel treated cohort^110^. However, we have to remark that no significant OS difference was found across BMI categories within the docetaxel treated patients. Similar mechanisms can be applied to the finding of a survival benefit for obese melanoma patients from the single included study. Several evidences indeed confirmed that a high BMI might be considered a surrogate marker of clinical benefit from immune checkpoint inhibitors for advanced melanoma patients^23 32 34^.

Interestingly, also obese RCC patients revealed to have a significantly longer OS compared to the respective non-obese cohorts. It has been already hypotesized that the perinephric white adipose tissue act as a reservoir of activated immune cells with increased characteristics of hypoxia, infiltration of Th1 cells, regulatory T cells, dendritic cells, and type 1 macrophages. However, only one out of six studies inclued patient receiving immunotherapy^23 52 36^.

Intriguingly, we found that the association between obesity and better clinical outcomes seems to be confirmed for those malignancies in which immune checkpoint inhibitors have first (and strongly) proved to be effective, despite studies involving patients receiving immune checkpoint inhibitors are poorly represented in this meta-analysis. Such results, might probably be an epiphenomenon, however we are allowed to speculate that the white adipose tissue could be considered an immune organ, which somehow plays a role in the anti-tumor immune response. It has been observed that the adipocyte-derived hormone leptin could alter T-cell function, resulting in improved response to anti-PD-1 therapy^11^. Moreover, another preclinical study reported that white adipose tissue acts as a reservoir for a peculiar population of memory T cells, which elicitate some effective responses in case of antigenic re-exposure during infections (and why not in case of exposure to cancer-specific antigens?)^2 37^.

Obese patients are also at increased risk of reduced physical activity. Various studies highlighted this concept. Physical activity declines over time in obese patients^23 82 39^. In particular, physical activity is strictly correlated with breast cancer and colorectal cancer mortality^240,241^. Therefore physical (in)activity should be a major target of obesity prevention and treatment in particular in cancer patients. Type 2 diabetes mellitus is strongly associated with obesity in the metabolic syndrome. More than 80 percent of cases of type 2 diabetes can be attributed to obesity, which may also account for many diabetes-related deaths. The association between BMI and cause-specific mortality was also illustrated in the Prospective Studies Collaboration analysis^242^. In the upper BMI range (25 to 50 kg/m2), each 5 kg/m^2^ increase in BMI was associated with a significant increase in mortality from coronary heart disease (CHD), stroke, diabetes mellitus, chronic kidney disease, and many cancers. In the same analysis, subjects with BMI below 22.5 kg/m2 had higher mortality compared with subjects with a BMI of 22.5 to 25 kg/m2. The excess mortality was predominantly due to smoking-related diseases (respiratory and cancer). Despite there are no clear recommendation about dosing of chemotherapy in obese patients, caution is recommended for high risk regimens^243^. The hypothesis that a reduced dose according to ideal body weight may lead to a reduced outcome cannot be confirmed by prospective studies but may be considered as potential reason for the observed results in some settings (e.g. breast cancer). In a pooled analysis of toxicity in obese vs non-obese rates of toxic effects were similar or lower in obese patients^244^.

However, this meta‐analysis has several limitations. First, we combined data of obese patients and compared prognosis with different non-obese subjects (normal weight or normal weight + overweight). Second, accurate measures of weight and height are always a challenge in observational studies. The evaluation is often before diagnosis but in some studies timing of obesity diagnosis is not described. Obese subjects have a general poor prognosis in term of overall mortality and non-cancer mortality, so it seems obvious that their prognosis is dismal. However, almost all studies provided a multivariate analysis according to main prognostic factor for oncological outcome so that obesity remain generally an independent prognostic factor in cancer patients. Duration of follow up, treatments received, and countries are heterogeneous even if subgroup analysis does not explain results with these different variables. Lastly, this meta‐analysis compared mortality between patients belonging to a fixed category of obesity (>30 kg/m^2^) and thus we are not able to provide effect size per unit increment of kg/m2.

In conclusion, results of this meta-analysis confirm and reinforce the notion that obesity is a competing risk factor for overall and cancer specific mortality and relapse in various cancers treated with curative intent or for metastatic disease except for lung cancer and melanoma where it has an apparent protective effect (obesity paradox). It is advised that in the future, oncologists increase their efforts to manage patients into multidisciplinary teams for care and cure of both cancer and obesity. Improving lifestyle (e.g., physical activity, calories intake, care/prevention of cardiovascular complications), more intensive follow-ups of cancer in obese, and adequate dose of medical therapies are all proved measures that may improve prognosis of obese cancer patients.

## Data Availability

Not applicable

---

**Suppl. file 2:**
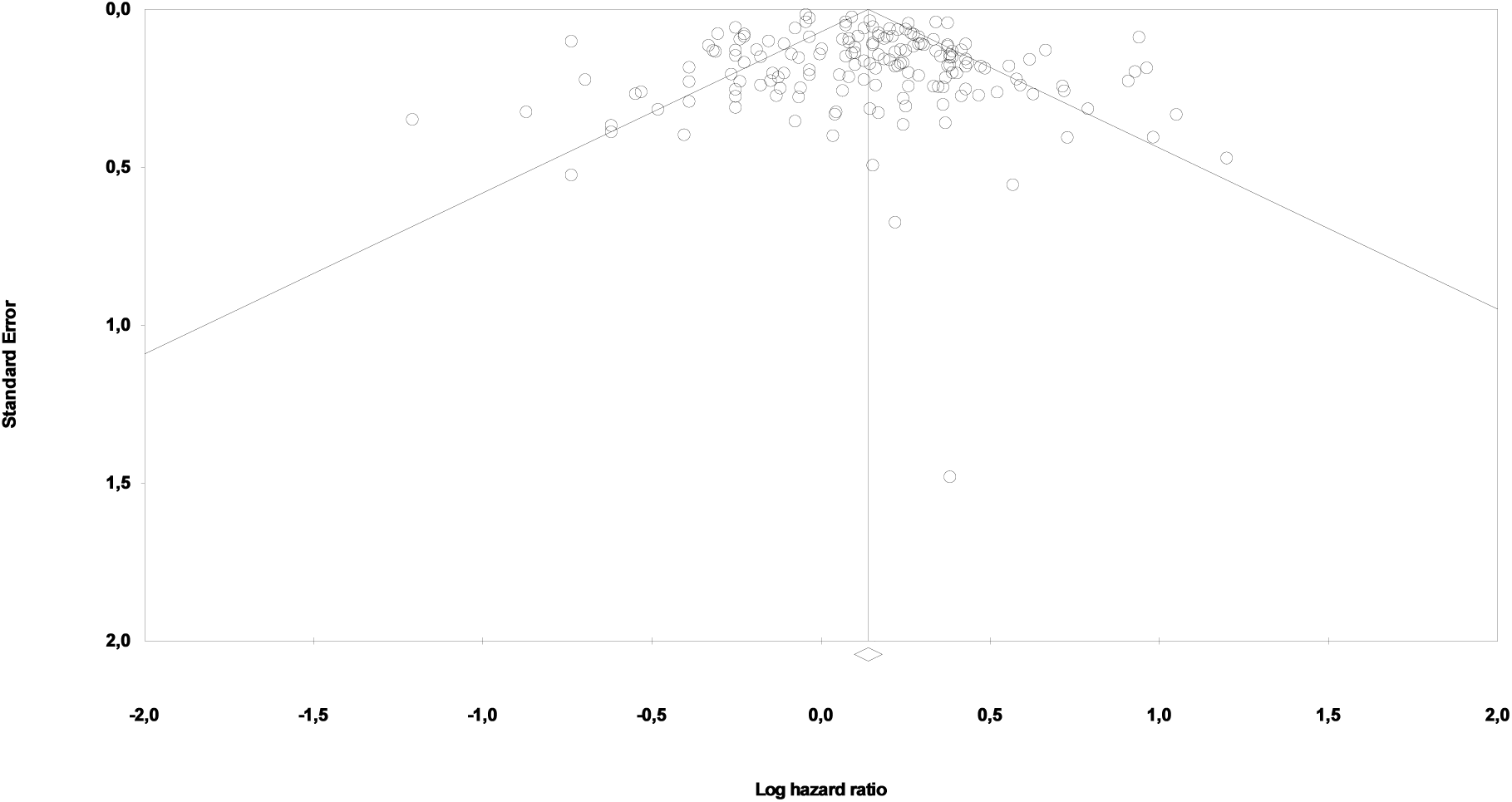
Funnel plot of publication bias for overall survival analysis

